# Beyond Accuracy: Multidimensional Evaluation of Large Language Models in Hepatocellular Carcinoma Management Emphasizing Prompting

**DOI:** 10.1101/2025.07.15.25331552

**Authors:** Jianchen Luo, Jing Ma, Tao Wang, Yiwen Qiu, Yi Yang, Haizhou Qiu, Guanhua Chen, Wentao Wang

**Author notes:** Trial registration number None.

## Abstract

**Background & Aims:** Hepatocellular carcinoma is the most common type of primary liver cancer and remains a major global health challenge. In resource-limited settings, patients often face barriers such as low screening rates, poor adherence, and limited access to medical information. Despite comprehensive clinical guidelines, issues like inadequate patient education and ineffective communication persist. While large language models show promise in clinical communication and decision support, their performance in hepatocellular carcinoma management has not been systematically evaluated across multiple dimensions.

**Methods:** Ten emerging language models, including general-purpose and medical-domain models, were assessed under prompted and unprompted conditions using a standardized question set covering five key stages: general knowledge, screening, diagnosis, treatment, and follow-up. Accuracy was rated by experts, while semantic consistency, local interpretability, information entropy, and readability were measured computationally.

**Results:** ChatGPT-4o and Grok-3 achieved the highest accuracy (2.62 ± 0.06, 93%; 2.60 ± 0.06, 95%) and interpretability (0.43;0.43). Prompting significantly improved accuracy (*p* < 0.001) and interpretability (*p* < 0.001) across all models. Semantic consistency declined slightly in most models; information entropy generally increased; readability changes varied.

**Conclusions:** This study presents the first multidimensional evaluation of large language models in hepatocellular carcinoma–related clinical tasks. General-purpose models outperformed some medical models, revealing limitations in domain-specific fine-tuning. Prompt design strongly influenced model performance. Further research should integrate diverse prompt strategies and clinical scenarios to improve the usability of language models in real-world oncology settings.

**Lay summary:** This study evaluated how well-advanced language-based artificial intelligence models can answer clinical questions related to hepatocellular carcinoma. The results showed that some models, especially when guided with structured instructions, provided accurate and understandable responses. These findings suggest that such tools may help improve communication and access to information for both doctors and patients managing liver cancer.

## Introduction

Hepatocellular carcinoma (HCC) is among the most common and lethal cancers worldwide. In 2022, approximately 865,300 new liver cancer cases were reported globally, accounting for 4.3% of all new cancers, with an estimated 757,900 deaths, representing 7.8% of cancer-related mortality^1^. HCC management involves a complex continuum, including screening, early diagnosis, treatment selection, and long-term follow-up^2^. Although international guidelines have standardized care pathways, real-world practice—especially in developing countries—still faces challenges such as delayed diagnosis, suboptimal treatment, and inconsistent follow-up^3^. Improving clinical decision support across diverse healthcare settings remains a critical issue in HCC care.

In recent years, large language models (LLMs), as a core technology in natural language processing, have shown great promise in medical applications^4^. Leading models like ChatGPT and Claude have performed at a level comparable to medical students on the United States Medical Licensing Examination (USMLE)^5^, and demonstrated potential in fields such as cardiology^6^ and surgery^7^. Domain-specific models have also shown strong transferability in tasks like Chinese clinical question answering and case summarization^8^. These advances suggest that LLMs could support clinical decision-making, medical education, and patient communication, especially in resource-limited settings^9^.

However, existing research on the medical capabilities of LLMs has predominantly focused on the accuracy of their responses^10^, with relatively limited attention given to the structural characteristics of model-generated responses. For instance, it remains unclear whether the information provided is focused and precise, whether it aligns with clinical guidelines, whether the text exhibits adequate readability and clinical applicability, and whether the model’s responses reflect interpretable semantic reasoning. These aspects are critical for evaluating the practical usability and deployment value of LLMs in clinical settings^11^.Therefore, a multidimensional quantitative framework is urgently needed to comprehensively analyze the response characteristics and potential advantages of LLMs in real-world medical tasks.

This study systematically evaluated the performance of LLMs in handling HCC-related clinical tasks under two conditions (with prompt and without prompt), using a standardized set of questions. It examined the variations in response quality, structural expression, and information organization, and explored the multi-dimensional impact of prompt information on model responses, providing quantitative insights for optimizing and applying LLMs in real-world clinical settings. The findings offer empirical references for clinicians in assessing and utilizing LLMs and provide interpretive support for improving model design and prompt strategies.

## Method

### Model selection and deployment

The language models evaluated in this study were selected based on the AlpacaEval leaderboard (updated on May 13, 2024), covering proprietary and open-source models, including several that are medically specialized. AlpacaEval is an automated evaluation framework developed by Stanford University to assess LLMs’ ability to respond to open-ended instructions across multiple tasks. The leaderboard ranks models based on pairwise preference comparisons with a strong reference model (e.g., GPT-4), reflecting their overall performance in terms of generation quality and utility^12^. Proprietary models were accessed via public or paid APIs, while open-source models were locally deployed on a high-performance consumer-grade workstation equipped with an Intel Core i7-13700KF processor, NVIDIA RTX 4090 GPU (24GB VRAM), and 64GB DDR5 6400MHz memory. All open-source models were quantized using 4-bit GPTQ to accommodate GPU memory limitations^13^. Since medically specialized models are underrepresented on the AlpacaEval leaderboard and lack systematic benchmarking, their inclusion was based on recent literature recommendations and GitHub popularity (i.e., number of stars)^14^.

### Question set and prompt construction

To systematically evaluate the response capabilities of LLMs in clinical tasks, we developed a structured question set covering key phases of HCC management, including general knowledge, screening, diagnosis and treatment, and follow-up. General knowledge questions were designed by clinical experts on our team. Screening-related questions were constructed based on the 2022 Guidelines for Liver Cancer Screening in the Chinese Population issued by the National Cancer Center of China (NCC-China)^15^ and the 2020 Guidelines for Stratified Screening and Monitoring of Primary Liver Cancer published by the Chinese Society of Hepatology and the Chinese Medical Association (CSH-CMA)^16^. Diagnosis, treatment, and follow-up questions referred to the 2024 Guidelines for the Diagnosis and Treatment of Primary Liver Cancer by the National Health Commission of China (NHC)^17^. To ensure contemporaneity, we also included the 2025 Hepatobiliary Cancers Clinical Practice Guidelines from the National Comprehensive Cancer Network (NCCN)^18^ and the 2025 Clinical Practice Guidelines for the Diagnosis, Treatment, and Follow-up of HCC issued by the European Society for Medical Oncology (ESMO)^19^. The question set primarily comprised closed-ended items, with a small proportion of open-ended questions included to evaluate model performance in more complex scenarios. Each question was paired with a predefined standard answer to ensure objectivity and reproducibility in the evaluation process.

In terms of prompt construction, this study adopted the CO-STAR framework^20^, which was recognized as a top-performing prompt design in the GPT-4 Prompt Engineering Competition. The framework consists of six elements: Context, Objective, Style, Tone, Audience, and Response format. To minimize variability in prompt formulation and ensure fair comparison across models, a standardized prompt was applied prior to each question.

### Question delivery and consistency analysis

Each question was input into each LLM three times to obtain three independent responses. Semantic Textual Similarity (STS) analysis was then performed to assess pairwise semantic similarity among the responses^21^ STS scores range from 0 (completely dissimilar) to 100 (completely identical), with a score above 90 considered semantically consistent. If all three responses were deemed semantically consistent, the original model was used to select the optimal answer. If semantic differences were detected, he response with the greatest overall semantic divergence was excluded based on STS scores, and the model was instructed to choose the best answer from the remaining two. This strategy aimed to mitigate generation instability and ensure the final evaluated response was both representative and consistent.

### Scoring procedure and criteria

Each response was independently and anonymously reviewed and rated by two clinical hepatology experts. To avoid subjective bias, the reviewers were blinded to both the model generating the response and whether a prompt was provided. If both experts agreed on a rating, it was directly adopted. If there was disagreement, they evaluated the response based on accuracy, adherence to clinical guidelines, clarity of communication, and practical usefulness to determine the final rating. According to previous studies, responses were classified into four levels: A (comprehensive and entirely correct), B (mostly correct, but with missing information or minor errors), C (contains major errors but includes some correct information), and D (completely incorrect or off-topic). The A/B/C/D ratings were assigned scores of 3, 2, 1, and 0, respectively.

### Local interpretability analysis

To evaluate the local interpretability of LLMs, this study employed the LIME (Local Interpretable Model-Agnostic Explanations) method to construct an explanation framework and conduct token-level semantic analysis of model-generated responses^22^. The number of samples for LIME was set to 10 times the number of tokens in the input sentence, following established settings in previous natural language processing tasks^23^. Based on this, Bio_ClinicalBERT was used to extract semantic features from each model response and identify key terms. These terms were then normalized through concept mapping (e.g., mapping “hepatocellular carcinoma” to “HCC”) and filtered by removing common English stopwords to minimize noise. The five terms with the highest absolute average weights across repeated runs were selected as the core explanatory terms for each response. For reference, clinical experts annotated five key terms for each question, and the overlap between expert- and model-identified terms was measured using the Intersection over Union (IoU). In addition, the interquartile range (IQR) of IoU values was calculated as a proxy for interpretability stability.

### Information entropy analysis

To complement conventional evaluation metrics for assessing the quality of responses, this study introduces information entropy as an auxiliary indicator to measure the determinacy and informational density of model-generated responses^24^. Entropy values were calculated for all responses from each model under both with prompt and without prompt conditions. The distribution patterns and variation trends were visualized to explore the impact of prompt information on LLM-generated responses.

### Readability analysis

To evaluate the expressive quality and comprehension difficulty of responses, this study introduced six commonly used English readability metrics: Flesch Reading Ease (FRE)^25^, Flesch-Kincaid Grade (FKG)^26^, Gunning Fog Index (GFI)^27^, Coleman-Liau Index (CLI)^28^, Automated Readability Index (ARI)^29^, and Dale-Chall Score (DCS)^30^. These metrics assess text readability from multiple dimensions, such as sentence length and word complexity. Based on these scores, we constructed a unified readability index, the Model Composite Readability Index (MCRI), for comprehensive quantitative assessment.

### Statistical Analysis

To enhance the robustness and comparability of various evaluation metrics, multiple statistical methods were employed throughout the analysis. Python (version 3.11) was used to implement STS, local interpretability, information entropy and readability analysis, along with data visualization. IBM SPSS Statistics (version 30.0.0) was used for statistical testing. The Friedman test was applied to evaluate overall differences across multiple groups. For paired comparisons, the Shapiro–Wilk test was first used to assess data normality. If data followed a normal distribution, paired t-tests were used; otherwise, the paired Wilcoxon Signed-Rank Test was applied, with multiple comparisons corrected using the False discovery rate (FDR) method. A p < 0.05 was considered statistically significant. For response quality assessment, mean scores and 95% confidence intervals (95% CI) were calculated. The combined proportion of responses rated as A or B was defined as the model’s effectiveness rate. In STS analysis, the proportion of semantically consistent responses among the three generated responses was defined as the model’s consistency rate. In the local interpretability analysis, each response was processed five times, and the extracted keywords and their weights were averaged to produce a stable explanation result. Higher IoU scores indicated greater overlap between expert- and model-identified terms, thus reflecting better local interpretability. For information entropy analysis, the mean entropy values with 95% CI were calculated for each model to evaluate uncertainty in responses. In the readability analysis, the 10% trimmed mean was computed for each model under each readability metric. These values were standardized using Z-score normalization to ensure consistent scale and comparability. Principal Component Analysis (PCA) was then conducted to integrate the six standardized readability scores. The first principal component (PC1) was extracted, and each model’s score along this dimension was defined as the composite readability index, MCRI. The loadings of each metric in PC1 represented their directional influence, while the normalized squared loadings (weight proportions) indicated each metric’s relative contribution to the overall readability structure. A higher MCRI score indicated lower overall readability, and vice versa.

## Result

### Model selection

A total of ten LLMs were included in this study. The selection criteria were based on AlpacaEval leaderboard performance, feasibility of local deployment, and adaptability to medical tasks. The proprietary models included ChatGPT-4o, Claude 3.5 Sonnet, ChatGLM4, DeepSeek-R1, Gemini2, Grok-3, Mistral Large, and HuatuoGPT II. The open-source models were Qwen2-7B-Instruct and BioMedLM 2.7B. Among them, HuatuoGPT II and BioMedLM 2.7B were specifically developed for medical applications. Some models were accessed via public web interfaces (either free or subscription-based), while others were used via API access or local deployment. Detailed characteristics of each model are presented in Table 1.

**Table 1.**
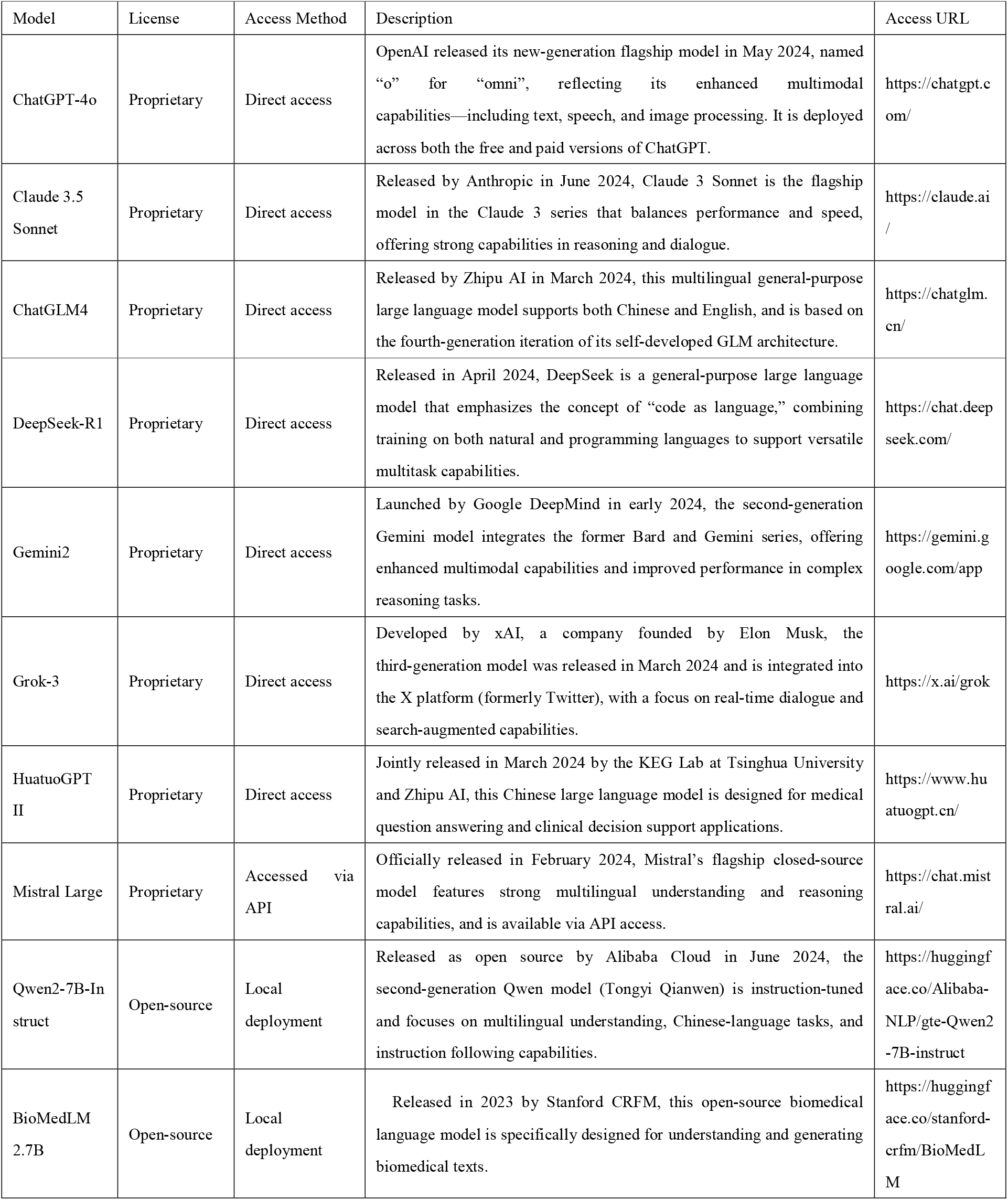
Characteristics of the models included in the study.

| Model | License | Access Method | Description | Access URL |
| --- | --- | --- | --- | --- |
| ChatGPT-4o | Proprietary | Direct access | OpenAI released its new-generation flagship model in May 2024, named “o” for “omni”, reflecting its enhanced multimodal capabilities—including text, speech, and image processing. It is deployed across both the free and paid versions of ChatGPT. | https://chatgpt.com/ |
| Claude 3.5 Sonnet | Proprietary | Direct access | Released by Anthropic in June 2024, Claude 3 Sonnet is the flagship model in the Claude 3 series that balances performance and speed, offering strong capabilities in reasoning and dialogue. | https://claude.ai/ |
| ChatGLM4 | Proprietary | Direct access | Released by Zhipu AI in March 2024, this multilingual general-purpose large language model supports both Chinese and English, and is based on the fourth-generation iteration of its self-developed GLM architecture. | https://chatglm.cn/ |
| DeepSeek-R1 | Proprietary | Direct access | Released in April 2024, DeepSeek is a general-purpose large language model that emphasizes the concept of “code as language,” combining training on both natural and programming languages to support versatile multitask capabilities. | https://chat.deepseek.com/ |
| Gemini2 | Proprietary | Direct access | Launched by Google DeepMind in early 2024, the second-generation Gemini model integrates the former Bard and Gemini series, offering enhanced multimodal capabilities and improved performance in complex reasoning tasks. | https://gemini.google.com/app |
| Grok-3 | Proprietary | Direct access | Developed by xAI, a company founded by Elon Musk, the third-generation model was released in March 2024 and is integrated into the X platform (formerly Twitter), with a focus on real-time dialogue and search-augmented capabilities. | https://x.ai/grok |
| HuatuoGPT II | Proprietary | Direct access | Jointly released in March 2024 by the KEG Lab at Tsinghua University and Zhipu AI, this Chinese large language model is designed for medical question answering and clinical decision support applications. | https://www.huatuogpt.cn/ |
| Mistral Large | Proprietary | Accessed via API | Officially released in February 2024, Mistral’s flagship closed-source model features strong multilingual understanding and reasoning capabilities, and is available via API access. | https://chat.mistral.ai/ |
| Qwen2-7B-Instruct | Open-source | Local deployment | Released as open source by Alibaba Cloud in June 2024, the second-generation Qwen model (Tongyi Qianwen) is instruction-tuned and focuses on multilingual understanding, Chinese-language tasks, and instruction following capabilities. | https://huggingface.co/Alibaba-NLP/gte-Qwen2-7B-instruct |
| BioMedLM 2.7B | Open-source | Local deployment | Released in 2023 by Stanford CRFM, this open-source biomedical language model is specifically designed for understanding and generating biomedical texts. | https://huggingface.co/stanford-crfm/BioMedLM |

### Response generation and consistency analysis

The evaluation consisted of 120 questions covering key clinical scenarios in the management of HCC, including 15 general knowledge questions, 19 screening questions, 11 diagnostic questions, 59 treatment-related questions, and 16 follow-up questions. This question set was designed to comprehensively assess the accuracy and guideline adherence of LLMs in supporting clinical decision-making for HCC, offering high medical representativeness and evaluation value (a complete list of questions is provided in Supporting Information 1). Under the “with prompt” condition, we employed structured prompts based on the CO-STAR framework to standardize the logical flow of responses and enhance clinical expression. The full prompt content is detailed in Supporting Information 2.

The consistency of responses was evaluated using STS analysis (responses and result of STS analysis are provided in Supporting Information 3). The proportion of response triplets exhibiting semantic equivalence was defined as the consistency rate for each model. Most LLMs demonstrated a decrease in response consistency under the with prompt condition. However, ChatGLM4 and Mistral Large were notable exceptions, showing improved consistency when prompts were provided. Although ChatGPT-4o exhibited a slight decrease in consistency, it maintained a high level under both with prompt and without prompt conditions. In contrast, HuatuoGPT II, BioMedLM 2.7B, and Qwen2-7B-Instruct consistently exhibited lower overall consistency rates (Table 2).

**Table 2.**
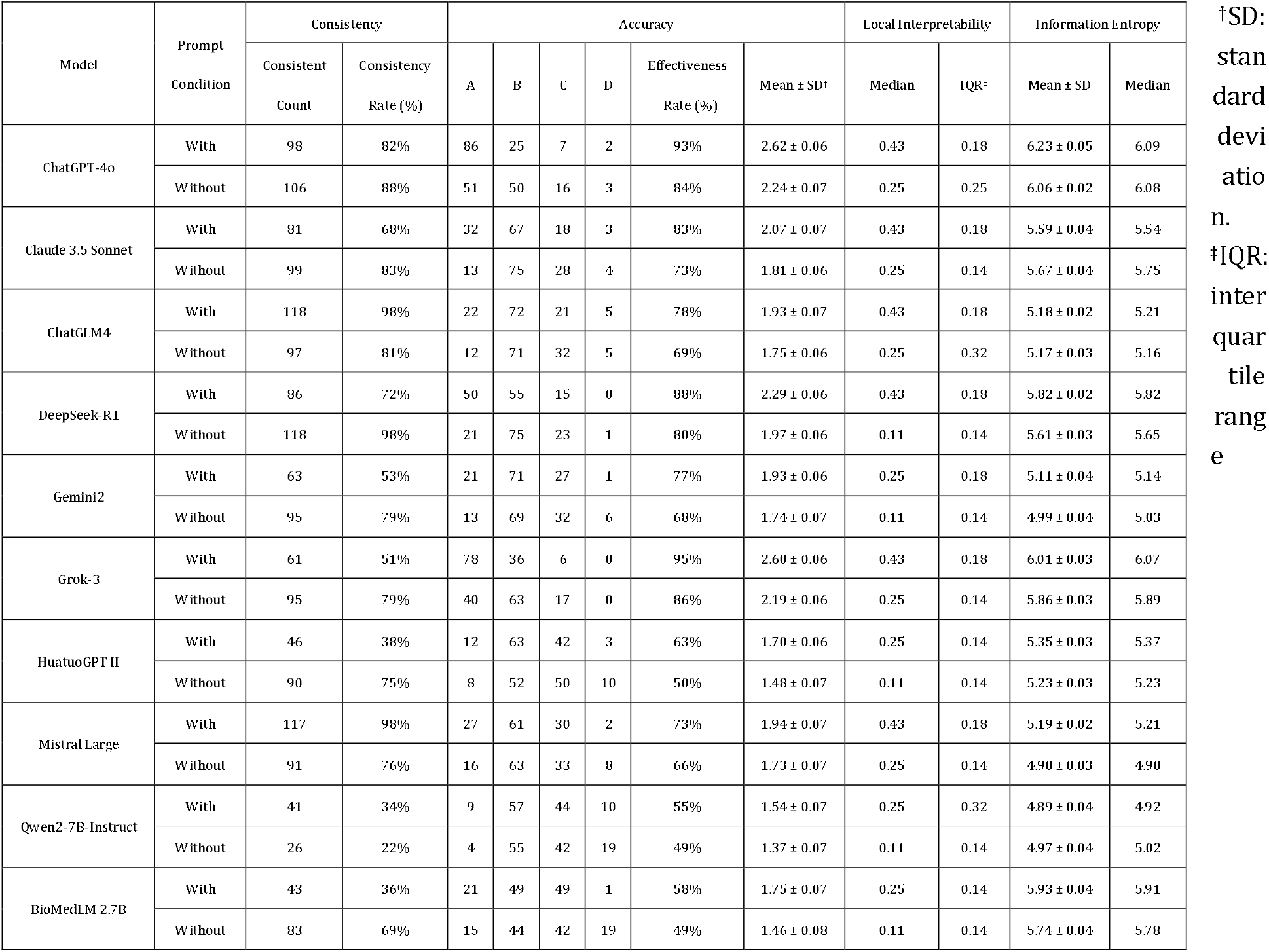
Stability, accuracy, local interpretability, and information entropy analysis results of models with and without prompt.

### Accuracy analysis

In the analysis of response accuracy, all models demonstrated higher average scores and effectiveness rates under the with prompt condition compared to without prompt. The best-performing models were ChatGPT-4o and Grok-3. Specifically, ChatGPT-4o yielded an average score of 2.62 ± 0.06 and an effectiveness rate of 93% under the with prompt condition, while under the without prompt condition, its average score was 2.24 ± 0.07 with an effectiveness rate of 84%. Grok-3 achieved an average score of 2.60±0.06 and an effectiveness rate of 95% with prompts, and 2.19 ± 0.06 with an effectiveness rate of 86% without prompts (Table 2). Both models consistently ranked at the top under both conditions. In contrast, the two medically specialized models—HuatuoGPT II and BioMedLM 2.7B—consistently ranked in the lower tier under both conditions. For HuatuoGPT II, the average score was 1.70 ± 0.06 with an effectiveness rate of 63% under the with prompt condition, and 1.48 ± 0.07 with an effectiveness rate of 50% without prompts. BioMedLM 2.7B scored an average of 1.75 ± 0.07 with an effectiveness rate of 58% under the with prompt condition, and 1.46 ± 0.08 with an effectiveness rate of 49% under the without prompt condition (Table 2). Qwen2-7B-Instruct recorded the lowest average scores and effectiveness rates among all models under both conditions. To provide a more intuitive visualization of the overall performance of different models, we developed a spiral-shaped graphic representation termed the “Nautilus Chart” (Figure 1). In this chart, the radius length and color intensity of each spiral segment correspond to the ranking of each model, highlighting their relative positions and differences in the accuracy evaluation. This visualization conveys a high density of information while offering strong visual clarity and differentiation. The code is provided in Supporting Information 4.

**Figure 1.** Accuracy score and efficiency ranking of model responses with and without prompt. a). Efficiency ranking of model responses with and without prompt. b). Accuracy score ranking of model responses with and without prompt.

The score data for each model did not follow a normal distribution (*p* < 0.01). Friedman tests were used to assess the overall differences among LLMs across paired tasks, revealing significant differences under both the with prompt and without prompt conditions (*p* < 0.001). Subsequently, pairwise comparisons between models were conducted. Under the with prompt condition, both ChatGPT-4o and Grok-3 showed significant differences compared to all other models (*p* < 0.001), but no significant difference was observed between the two (*p* = 0.78). DeepSeek-R1, which ranked slightly lower, also differed significantly from most other models (*p* < 0.01). The two medically specialized models, HuatuoGPT II and BioMedLM 2.7B, significantly differed from the majority of other models (*p* < 0.05), but not from each other (*p* = 0.63). Qwen2-7B-Instruct performed significantly worse than all other models (*p* < 0.05). No significant differences were observed among ChatGLM4, Gemini2, Claude 3.5 Sonnet, and Mistral Large. Under the without prompt condition, ChatGPT-4o and Grok-3 again significantly outperformed other models (*p* < 0.001), with no significant difference between them (*p* = 0.54). DeepSeek-R1 remained significantly different from most models (*p* < 0.01), though the difference with Claude 3.5 Sonnet was marginally insignificant (*p* = 0.05). HuatuoGPT II and BioMedLM 2.7B continued to show significant differences from most models (*p* < 0.01), but no significant difference between each other (*p* = 0.76), and their differences with Qwen2-7B-Instruct were no longer significant (*p* = 0.11, *p* = 0.24). No significant differences were observed among ChatGLM4, Gemini2, Claude 3.5 Sonnet, and Mistral Large (Table 2). All models scored significantly higher under the with prompt condition compared to the without prompt condition (Figure 2).

**Figure 2.** Pairwise significance analysis comparing model performance with and without prompt. The upper triangle shows the significance levels between models under the with prompt condition, while the lower triangle presents the significance levels under the without prompt condition. The diagonal section indicates the significance of performance differences within each model between the with prompt and without prompt conditions. ^*^ *p* < 0.05, ^**^ *p* < 0.01, ^***^ *p* < 0.001.

### Local interpretability analysis

The IoU for all models did not follow a normal distribution (*p* < 0.01). Therefore, the Wilcoxon signed-rank test was used to evaluate differences between the with prompt and without prompt conditions. Results showed statistically significant differences across all models between the two conditions (*p* < 0.05 for HuatuoGPT II, and *p* < 0.001 for all other models) (Figure 3). Under the with prompt condition, the median IoU was 0.43 for ChatGPT-4o, Claude 3.5 Sonnet, ChatGLM4, DeepSeek-R1, Grok-3, and Mistral Large, while the remaining four models had a median IoU of 0.25. Notably, Qwen2-7B-Instruct showed the greatest dispersion (IQR = 0.32), whereas the two medically specialized models, HuatuoGPT II and BioMedLM 2.7B, exhibited more concentrated distributions (IQR = 0.14). Under the without prompt condition, the median IoU was 0.25 for ChatGPT-4o, Claude 3.5 Sonnet, ChatGLM4, Grok-3, and Mistral Large, and 0.11 for the other five models. ChatGLM4 demonstrated the highest level of dispersion (IQR = 0.32), while most other models had relatively low IQR values (approximately 0.14); the IQR of ChatGPT-4o was 0.25. Of particular note, DeepSeek-R1 exhibited the largest change in median IoU between the without prompt and with prompt conditions (0.11 vs. 0.43) (Table 2). The extracted keywords from all model responses and their IoU are provided in Supporting Information 5.

**Figure 3.** Local interpretability analysis by intersection over union for included models with or without prompt.

### Information entropy analysis

In the analysis of information entropy, most models demonstrated increased mean and median entropy values under the with prompt condition (Table 2). Among them, Mistral Large exhibited the largest increase, with a median entropy of 5.19 ± 0.02 under the with prompt condition, compared to 4.90 ± 0.03 under the without prompt condition. In contrast, Claude 3.5 Sonnet and Qwen2-7B-Instruct showed slight decreases in both average and median entropy following prompt inclusion. Specifically, the median entropy for Claude 3.5 Sonnet decreased from 5.67 ± 0.04 to 5.59 ± 0.04, and for Qwen2-7B-Instruct, it decreased from 4.97 ± 0.04 to 4.89 ± 0.04. Statistical tests showed that all models except Claude (*p* = 0.10) and ChatGLM4 (*p* = 0.95) exhibited significant differences in entropy between the with prompt and without prompt conditions. Specifically, Qwen2-7B-Instruct showed *p* < 0.05, ChatGPT-4o showed *p* < 0.01, and the remaining six models showed *p* < 0.001(Figure 4). Information entropy of all model responses provided in Supporting Information 6.

**Figure 4.** Distribution of information entropy for each model’s responses under both with prompt and without prompt conditions. a). The distribution of information entropy for responses of ChatGPT-4o, b). The distribution of information entropy for responses of Claude 3.5 Sonnet, c). The distribution of information entropy for responses of ChatGLM4, d). The distribution of information entropy for responses of DeepSeek-R1, e). The distribution of information entropy for responses of Gemini2, f). The distribution of information entropy for responses of Grok-3, g). The distribution of information entropy for responses of Mistral Large, h). The distribution of information entropy for responses of Qwen2-7B-Instruct, i). The distribution of information entropy for responses of HuatuoGPT II, j). The distribution of information entropy for responses of BiomedLM 2.7B.

### Readability analysis

In the readability analysis, we constructed a composite readability score—termed the MCRI—based on six standardized readability metrics: FRE, FKGL, CLI, GFI, ARI, and DCS. PCA was used for dimensionality reduction and score integration. The PCA results indicated the weight proportions of each metric in PC1 were as follows: FRE (18%), FKGL (20%), CLI (15%), GFI (19%), ARI (19%), and DCRS (9%), with the PC1 explaining approximately 80% of the total variance. The z-score standardized scores and raw values for each readability metric are presented in Table 3. Based on the MCRI, ChatGPT-4o, Claude 3.5 Sonnet, Grok-3, and Qwen2-7B-Instruct showed an increasing trend under the with prompt condition, while the remaining LLMs demonstrated a decreasing trend, with Mistral Large showing the most pronounced reduction. Notably, Claude 3.5 Sonnet exhibited relatively high MCRI values under both with prompt and without prompt conditions (Figure 5). The results of the readability analysis for each model’s responses are provided in Supporting Information 7.

**Table 3.** Readability analysis results and scores for each rating criterion and their weights.

| Model | Prompt Condition | Readability |  |  |  |  |  |  |
| --- | --- | --- | --- | --- | --- | --- | --- | --- |
|  |  | MCRI<sup>1</sup> | FRE<sup>2</sup>(18%)<br>Z-score (raw score) | FKG<sup>3</sup>(20%)<br>Z-score (raw score) | GFI<sup>4</sup>(19%)<br>Z-score (raw score) | CLI<sup>5</sup>(15%)<br>Z-score (raw score) | ARI<sup>6</sup>(19%)<br>Z-score (raw score) | DCS<sup>7</sup>(9%)<br>Z-score (raw score) |
| ChatGPT-4o | With | -1.21 | 0.57 (10.66) | -0.03 (18.31) | -0.37 (20.06) | -0.64 (17.40) | 0.04 (20.49) | -1.84 (11.90) |
|  | Without | -2.56 | 1.03 (14.62) | -0.73 (17.12) | -1.09 (18.69) | -0.94 (16.91) | -0.73 (18.96) | -2.12 (11.68) |
| Claude 3.5 Sonnet | With | 4.50 | -2.57 (-16.36) | 1.23 (20.46) | 1.21 (23.10) | 3.36 (24.00) | 1.32 (22.99) | 1.43 (14.51) |
|  | Without | 4.18 | -1.98 (-11.28) | 1.86 (21.54) | 1.57 (23.78) | 1.82 (21.47) | 2.05 (24.43) | 0.74 (13.96) |
| ChatGLM4 | With | -0.47 | 0.32 (8.54) | -0.03 (18.31) | 0.05 (20.88) | -0.46 (17.71) | -0.15 (20.12) | -0.31 (13.12) |
|  | Without | 0.44 | -0.18 (4.22) | 0.59 (19.37) | 0.45 (21.65) | -0.43 (17.76) | 0.39 (21.17) | -0.31 (13.12) |
| DeepSeek-R1 | With | -3.07 | 1.21 (16.20) | -1.50 (15.81) | -1.50 (17.90) | -0.74 (17.25) | -1.49 (17.47) | -1.02 (12.56) |
|  | Without | -1.69 | 0.44 (9.51) | -0.86 (16.89) | -0.85 (19.13) | -0.33 (17.92) | -1.06 (18.32) | -0.55 (12.93) |
| Gemini2 | With | -1.85 | 0.88 (13.34) | -0.85 (16.92) | -0.76 (19.32) | -0.80 (17.14) | -0.88 (18.67) | -0.24 (13.18) |
|  | Without | 0.01 | -0.34 (2.88) | 0.04 (18.43) | -0.19 (20.41) | 0.03 (18.51) | -0.18 (20.05) | -0.01 (13.36) |
| Grok-3 | With | -0.53 | 0.39 (9.13) | -0.65 (17.26) | -0.53 (19.75) | -0.04 (18.39) | -0.36 (19.69) | 1.07 (14.22) |
|  | Without | -1.32 | 0.66 (11.45) | -0.90 (16.83) | -0.79 (19.26) | -0.36 (17.87) | -0.74 (18.95) | 0.53 (13.79) |
| HuatuoGPT II | With | -1.81 | 1.03 (14.62) | -0.56 (17.40) | -0.51 (19.79) | -1.05 (16.73) | -0.52 (19.38) | -0.86 (12.69) |
|  | Without | -0.26 | 0.07 (6.35) | 0.30 (18.88) | 0.16 (21.09) | -0.59 (17.50) | 0.02 (20.44) | -0.71 (12.80) |
| Mistral Large | With | -1.08 | 0.41 (9.32) | -0.48 (17.54) | -0.37 (20.06) | -0.25 (18.06) | -0.57 (19.27) | -0.62 (12.88) |
|  | Without | 4.34 | -1.55 (-7.59) | 2.46 (22.55) | 2.45 (25.48) | 0.61 (19.46) | 2.45 (25.22) | 0.79 (14.00) |
| Qwen2-7B-Instruct | With | 0.55 | 0.11 (6.70) | 0.06 (18.47) | 0.56 (21.84) | -0.06 (18.37) | 0.12 (20.64) | 0.98 (14.15) |
|  | Without | 2.77 | -1.01 (-2.93) | 1.30 (20.58) | 1.62 (23.88) | 0.57 (19.40) | 1.20 (22.75) | 1.07 (14.22) |
| BioMedLM 2.7B | With | -0.11 | 0.17 (7.24) | -0.46 (17.58) | -0.35 (20.10) | 0.11 (18.65) | -0.27 (19.88) | 1.28 (14.39) |
|  | Without | -0.81 | 0.34 (8.73) | -0.79 (17.02) | -0.76 (19.31) | 0.19 (18.78) | -0.64 (19.14) | 0.73 (13.95) |

<sup>1</sup>MCRI: model composite readability index.
<sup>2</sup>FRE: Flesch reading ease.
<sup>3</sup>FKG: Flesch-Kincaid grade.
<sup>4</sup>GFI: Gunning Fog index.
<sup>5</sup>CLI: Coleman-Liau index.
<sup>6</sup>ARI: automated readability index.
<sup>7</sup>DCS: Dale-Chall score

**Figure 5.** Readability analysis results of each model’s responses under both with prompt and without prompt conditions. The MCRI values and trend of each model’s responses under both with prompt and without prompt conditions. The dashed lines divide the chart into four quadrants: the upper left quadrant represents high readability with high scores, the lower left quadrant represents high readability with low scores, the upper right quadrant represents low readability with high scores, and the lower right quadrant represents low readability with low scores. MCRI: model composite readability index.

## Discussion

### Principal results

This study systematically evaluated multiple mainstream large language models in key clinical tasks related to hepatocellular carcinoma, including general knowledge, screening, diagnosis, treatment, and follow-up. Models such as ChatGPT-4o and Grok-3 showed strong clinical applicability in response accuracy and language structure, with several findings reported for the first time. Notably, this is the first study to assess LLMs multidimensionally through local interpretability, information entropy, and readability, offering new insights into response quality and controllability. Prompt information was embedded into the design using a structured CO-STAR framework, demonstrating for the first time that prompts influence not only accuracy but also semantic focus, density, and linguistic structure. This provides a methodological reference for future prompt optimization. Additionally, we introduced a composite readability index (MCRI) and a novel visualization format (Nautilus Chart) to support more interpretable evaluations of model responses in clinical contexts.

### Comparison with prior work

In the semantic consistency analysis, most models showed decreased scores under prompted conditions, suggesting that prompts may introduce more diverse linguistic expression and increase response variability. In contrast, ChatGLM4 and Mistral Large showed improved consistency after prompting, indicating more stable and focused responses. ChatGPT-4o maintained high consistency in both conditions, demonstrating strong generative stability, consistent with prior findings by van Nuland M et al.^31^. HuatuoGPT II, BioMedLM 2.7B, and Qwen2-7B-Instruct scored lower overall, reflecting limited stability and controllability, which may affect their reliability in clinical use.

The accuracy analysis revealed that ChatGPT-4o and Grok-3 achieved high average scores and validity rates under both prompt conditions, indicating strong medical knowledge coverage and response accuracy. These findings are consistent with previous reports on the ChatGPT series by Son M et al.^32^, while the performance of Grok-3 is reported here for the first time. By contrast, the medically specialized models HuatuoGPT II and BioMedLM 2.7B obtained relatively lower scores, suggesting limitations in knowledge depth or reasoning capability when dealing with open-ended clinical tasks, as similarly reported by Zaghir J et al.^33^. Qwen2-7B-Instruct ranked lowest in both conditions, implying that despite its large model size, it may lack adequate generalizability in English medical tasks. Prompting significantly improved response accuracy across all models, with the effect being particularly notable in moderately performing models, further underscoring the importance of prompt design in enhancing model performance and clinical applicability.

Local interpretability refers to whether a model’s response to a single input is understandable and grounded in key input information^34^. In medical tasks, this is critical for response credibility, clinical usability, and regulatory acceptance^35^. We used the LIME method to extract high-weight terms from responses and compared them to expert-annotated keywords, calculating IoU as a measure of interpretability. ChatGPT-4o, Claude 3.5 Sonnet, ChatGLM4, Grok-3, and Mistral Large showed higher IoU scores under both conditions, indicating better focus on key information. All models improved significantly with prompts, suggesting a universal benefit. Notably, ChatGPT-4o and ChatGLM4 showed more consistent semantic focus when prompted, aligning with previous findings by Kuerbanjiang W et al.^36^.

This study is the first to examine how prompt information affects the entropy of responses generated by large language models. Most models showed significantly higher entropy with prompts, indicating more information-dense and complex outputs^37^. Mistral Large had the greatest increase, while Claude 3.5 Sonnet and Qwen2-7B-Instruct showed slight decreases, suggesting weaker prompt responsiveness or stable generation styles. All models except Claude and ChatGLM4 showed significant entropy differences. As entropy reflects content richness or focus—not quality per se—these findings highlight varying strategies in information organization across models^38^.

To assess the practical readability of language model responses in clinical communication, this study integrated six established English readability metrics into a composite index (MCRI) via principal component analysis. A higher MCRI score reflects lower readability. The SMOG^39^ index was excluded due to its requirement for texts with at least 30 sentences, which many responses did not meet^40^. Results showed that most models had lower MCRI scores with prompts, suggesting improved readability. Mistral Large showed the greatest improvement, while models such as ChatGPT-4o, Claude 3.5 Sonnet, and Qwen2-7B-Instruct showed slight increases, potentially due to more complex or denser responses. Claude 3.5 Sonnet consistently produced higher MCRI scores, indicating a trade-off between clarity and professionalism. Overall, prompt input improved readability in most models, although the effects varied.

In summary, this study is the first to integrate semantic consistency, accuracy, interpretability, entropy, and readability into a comprehensive evaluation framework for LLMs in medical tasks. It provides a more holistic approach compared to prior studies focused on accuracy or pass rates, such as USMLE simulations^5^ or simplified guidelines^9^. Results showed general-purpose models like ChatGPT-4o and-3 demonstrated strong adaptability, aligning with studies on large model generalization^41^. Notably, this is the first to report Grok-3’s performance in HCC-related contexts. Medically specialized models (e.g., HuatuoGPT II, BioMedLM 2.7B) showed limitations in open-ended tasks, challenging the idea that fine-tuned models should outperform general-purpose ones^42^. Specialized models focus on structured knowledge recall, limiting their flexibility in complex tasks. In contrast, general-purpose models excel in contextual understanding, particularly with prompt guidance. Medical models often lack instruction tuning, limiting their response to complex prompts^43^. Additionally, their narrow domain training limits generalizability. These findings suggest that medical LLMs should focus not only on domain knowledge but also on instruction alignment, language generation, and multitask adaptability^44^. The weaker performance of Qwen2-7B-Instruct may be due to its optimization for Chinese-language contexts and local computational constraints^45^.

This study found that under the with prompt condition, all models improved response accuracy, and local interpretability generally increased, suggesting that prompt information helps models focus on key information, enhancing task execution in medical contexts^46^. However, trends in information entropy and readability varied across models. For example, Mistral Large generated more detailed responses with increased entropy and decreased readability, while DeepSeek-R1 improved response quality while maintaining linguistic conciseness. These findings suggest that models adopt different strategies to improve accuracy—some by expanding information coverage, others by enhancing semantic focus^47^. This underscores the importance of tailored prompt strategies for optimal semantic control and task alignment.

### limitations

This study systematically evaluated mainstream LLMs in HCC-related tasks but has several limitations. First, while the question set covers key clinical processes (screening, diagnosis, treatment, and follow-up), it does not address all disease subtypes or individualized treatment scenarios, limiting model adaptability. Second, response quality was rated by two hepatology experts, which, despite blinding, may still introduce some subjectivity. Additionally, metrics like information entropy and readability quantify response structure but require further validation regarding their link to clinical cognitive load and reading experience^48^. Finally, the standardized prompt template used did not explore the effects of varied prompt styles, and real-world user interaction was not considered. Future research should incorporate diverse prompt strategies and real clinical scenarios to improve external validity and practical applicability.

### Future directions

As LLMs evolve in healthcare, their potential in decision support, patient education, and intelligent question answering grows. This study shows that prompt information improves accuracy and influences model responses, such as language structure and semantic focus. Prompt engineering optimizes performance and regulates the model’s “language style”^49^. Future research should explore customized prompt strategies for diverse user groups (e.g., physician vs. patient) to enhance adaptability. These strategies may enable context-aware AI systems for real-world applications like patient communication^50^. Future work could also integrate multimodal inputs (e.g., health records, imaging) with LLMs for complex clinical decision support. Ethical concerns on accessibility and fairness should ensure AI tools are accessible and interpretable^50^.

## Data Availability

None

## Conclusions

This study proposed a multidimensional framework—including accuracy, consistency, interpretability, entropy, and readability—to evaluate mainstream LLMs in HCC-related clinical tasks. Models like ChatGPT-4o and Grok-3 showed strong performance and clinical potential. Prompting improved accuracy and influenced response structure and style, underscoring its role in guiding model behavior. These results support standardized evaluation and prompt design in medical LLM deployment. Future research should incorporate clinical interactions, user feedback, and multimodal data to develop more intelligent and personalized tools.

### Abbreviations

HCC: hepatocellular carcinoma
LLMs: large language models
USMLE: United States Medical Licensing Examination
NCC-China: National Cancer Center of China
CSH-CMA: Chinese Society of Hepatology and the Chinese Medical Association
NHC: National Health Commission of China
NCCN: National Comprehensive Cancer Network
ESMO: European Society for Medical Oncology
STS: semantic textual similarity
LIME: local interpretable model-agnostic explanations
IoU: intersection over union
IQR: interquartile range
FRE: Flesch reading ease
FKG: Flesch-Kincaid grade
GFI: Gunning Fog index
CLI: Coleman-Liau index
ARI: automated readability index
DCS: Dale-Chall score
MCRI: model composite readability index
PCA: principal component analysis
PC1: the first principal component
FDR: false discovery rate

## Acknowledgments

We would like to sincerely thank the Clinical Epidemiology and Evidence-Based Medicine Center for their valuable contributions to this study. Special thanks to Xin Zhang for her valuable feedback and assistance in refining the statistical methods.

